# Viable Monkeypox virus in the environment of a patient room

**DOI:** 10.1101/2022.09.15.22280012

**Authors:** Kalisvar Marimuthu, Judith Chui Ching Wong, Poh Lian Lim, Sophie Octavia, Xiaowei Huan, Yi Kai Ng, Jun Jing Yang, Stephanie Sutjipto, Kyaw Zaw Linn, Yin Xiang Setoh, Chong Hui Clara Ong, Jane Griffiths, Sharifah Farhanah, Thai Shawn Cheok, Nur Ashikin Binti Sulaiman, Sipaco Barbara Congcong, Erica Sena Neves, Liang Hui Loo, Luqman Hakim, Shuzhen Sim, Merrill Lim, Mohammad Nazeem, Shawn Vasoo, Kwok Wai Tham, Oon Tek Ng, Lee Ching Ng

**Author notes:** **Corresponding Author 1** Kalisvar Marimuthu, Senior Consultant, Address: National Centre for Infectious Diseases, 16 Jalan Tan Tock Seng, Singapore 308442., **Corresponding Author 2** Lee Ching Ng, Environmental Health Institute, National Environment Agency, 11 Biopolis Way, Helios Block, #06-05/08, Singapore 138667. These authors contributed equally (Kalisvar Marimuthu, Judith Chui Ching Wong).

## Abstract

We conducted a prospective environmental surveillance study to investigate the air, surface, dust and water contamination of a room occupied by a patient infected with Monkeypox virus (MPXV) at various stages of his illness. The patient tested positive for MPXV from a throat swab and skin lesions. Environmental sampling was conducted in a negative pressure room with 12 unidirectional HEPA air changes per hour and daily cleaning of the surfaces. A total of 179 environmental samples were collected on days 7, 8, 13, and 21 of his illness. Air, surface, and dust contamination was highest during the first eight days of the illness, with a gradual decline to the lowest contamination level by day 21. Viable MPXV was isolated from surfaces and dust samples and no viable virus was isolated from the air and water samples.

**Synopsis:** Inadequate research prevail on the mode of transmission for monkeypox virus. This study reports the findings of viable monkeypox virus from the patient environment, highlighting its implications for human health and impact on infection and prevention control measures.

## Introduction

The multicountry Monkeypox virus (MPXV) outbreak of 2022 continues to expand. As of 22 July 2022, more than 16,000 laboratory-confirmed cases, including five deaths, have been reported from 75 countries in five World Health Organization (WHO) regions, leading the WHO Director General to declare the Monkeypox outbreak a global health emergency^1^. Since the first reported case of human infection in 1970, the MPXV has caused multiple outbreaks in traditionally endemic regions of central and western Africa, with a small number of imported cases and limited transmission documented outside these areas^2^. With the decrease in herd immunity provided by Smallpox vaccination, human encroachment into the forested area, and increased global mobility, the epidemiology of MPXV has changed over the years with increased cases^2^ and secondary attack rates^3^.

Wild animal contact in traditionally endemic areas and close physical contact with infected individuals are known risk factors for MPXV human infections^3^. Fomite-based transmission through contaminated linen and rooms occupied by infected patients has also previously been shown^4^. Epidemiological evidence suggests that the current 2022 outbreak is fuelled by direct physical contact^5^, though a reported prolonged throat positivity for MPXV even after the resolution of skin lesions^6^, raised the concern for aerosol-based transmissions. Other possible modes of human-to-human transmission of MPXV have not been systematically investigated, to fully understand the transmission dynamics of MPXV. This study aims to investigate the extent of viral contamination of the air, surface, dust and water environment of a room occupied by a patient with MPXV at various stages of illness.

## Methods

### Study design, setting and patient selection

We conducted longitudinal sampling of air, surfaces, water and dust in the room occupied by the MPXV patient. Environmental sampling was done on days 7, 8, 13, and 21 of illness. An additional air sampling was conducted on day 15 of the illness. Supplementary Figure 1 illustrates the locations of environmental sampling. The patient was cared for in an airborne infection isolation room (AIIR) at the National Centre for Infectious Diseases (NCID), Singapore. The AIIR room had 12 air changes/h, an average temperature of 23□C, relative humidity of 52-60% and an exhaust flow of 570 m^3^/h. The floor and high-touch surfaces in the patient’s room and toilet floor were cleaned daily with bleach 10,000 ppm. On the day of environmental sampling, the room cleaning was done after the completion of sampling. The patient was on the bed during the sampling except for 5 minutes when the study team needed to collect vacuumed dust samples from the linen.

### Air sampling

We sampled the patient’s room air using SASS3100 (Research International) and Coriolis μ (Bertin Instruments) samplers (Supplementary Table 1). On days 7 and 8 of illness, two SASS and Coriolis samplers were placed on the left and right sides of the patient. The air samplers were set at 0.8 meters on the left and 0.9 meters on the patient’s right, and they were positioned on a trolley at 1.2 meters from the ground. On days 13 and 21 of illness, an additional set of SASS and Coriolis air samplers were placed at 2.5 meters from the patient at 1.2 meters from the ground. The SASS samplers were run for 2 h at a flow rate of 300 L/min, and the Coriolis sampler was run at 100 L/min for 1.5 h and collected in 2-5 mL of viral transport medium (VTM). A control sample with the air samplers turned off was collected immediately before the SASS and Coriolis samplers were turned on. Particle size measurements were concurrently collected on days 7 and 8 using the DustTrak™ DRX 8534. The DustTrak™ DRX 8534 was placed next to one set of SASS and Coriolis samplers on the left side of the patient Measurements were taken every min for a duration of 0.25 hr before sampling, and a duration of 2h when sampling was carried out.The concentration level of particle size of PM1, PM 2.5, PM4, PM 10 were collected.

On day 15 of illness, additional air samples were collected with 4 NIOSH BC 251 bioaerosol and two SASS samplers. NIOSH samplers were connected to SKC Universal air sampling pumps set at a flowrate of 3.5 L/min and run for 4 h, collecting three different size ranges of particles: less than 1 μm, 1-4 μm, more than 4 μm). The NIOSH samplers were placed 0.8 meters on the left and 0.9 meters on the patient’s right at the height of 1.2 meters from the ground.

### Surface sampling

We used sterile nylon flocked swabs (Puritan UniTranz-RT, Puritan Medical Products) to collect surface samples. Swabs were moistened with universal transport medium before surfaces were sampled. We collected surface swab samples from the patient’s room (floor, overbed table, bed rails, control panels, call bell, bedside locker, patient’s chair, switched over the bed, and stethoscope, personal protective equipment racks, external and internal surfaces of the sink, glass window panel, and sliding door), toilet (toilet door handle, toilet seat, hand support rails, and external and internal surfaces of the sink), anteroom (floor, sliding door to the room, sliding door to the clean corridor, external and internal surfaces of the sink)

### Dust sampling

Dust samples from linen, room and toilet floor surfaces were collected using vacuum socks (X-Cell-200, Midwest Filtration) fitted on a vacuum pump (Omega Plus HEPA Abatement Vacuum, Atrix international). After sampling, the socks were folded and detached from the vacuum pump. Zip-tie was used to close the open end of the collected dust sample and the entire sock was placed into a sterile Ziplock bag and transported back to the lab.

### Water sampling

We collected water samples from the P-trap of sinks in the patient’s room and toilet on days 7, 8, 13, and 21. Nasogastric feeding tubes connected to 50 ml syringes were used to collect 20-30 mls of water from P-trap, which were then transported to the lab in hospital urine collection cups.

### Quality control during sampling

Two study team members collected samples while wearing N95 respirators, faces shield, hair cover, water-resistant full sleeve gowns, gloves, and shoe covers. Two research assistants observed the process and ensured adherence to the environmental sampling protocol.

### Sample transfer and processing

All samples were immediately placed in a cooler box with ice packs and transferred to the biosafety level-3 (BSL-3) laboratory at the Environmental Health Institute, National Environmental Agency of Singapore following prevailing biosafety guidelines.

#### Laboratory methods

##### Sample processing

SASS samples were processed following the protocol adopted from^13^ Briefly, the filter membrane was soaked in a tube containing□1 mL of phosphate-buffered saline with 0.1% (v/v) Triton X-100 and incubated for 30 min. A sterile syringe was then used to retrieve the suspension solution from the membrane for DNA extraction. Coriolis samples collected in VTM were directly used for viral DNA extraction and virus culture. For NIOSH samples, particles collected on sample tubes and filters were resuspended with 1 mL M199 media, vortexed and used for viral DNA extraction and culture.

Fomite swabs collected in UTM were vortexed at high speed for 60 secs. The swabs were carefully removed and the sample was directly used for viral DNA extraction and virus culture.

Dust samples were processed in two ways. For selected samples, dust and visible scabs collected in vacuum socks were swabbed using a premoistened swab. The swab was resuspended in 1 mL phosphate-buffered saline (PBS). The vacuum sock was then retained in an environmental chamber to assess virus stability over time.

Water samples were processed using Amicon ultrafiltration centrifugal devices following the protocol described by Mailepessov *et al*.^14^.

### DNA extraction and quantitative Real-Time PCR

DNA extraction was carried out using the Qiagen DNeasy Blood and Tissue kit. A pre-extraction inactivation step was carried out where 200 μL of sample was added to 200 μL of Qiagen Buffer AL and 20 μL of proteinase K for sample lysis and inactivation at 56±2°C for 30 min. The manufacturer’s protocol was subsequently followed for DNA extraction (QIAGEN, Germany).

Real-Time PCR was conducted on a Quantstudio Real-Time PCR machine (Thermofisher Scientific, USA) using specific primers targeting the Monkey Pox B7 gene previously described^15^. The reaction mixture contains 10μL of LUNA™ (Roche, Switzerland) RT-Probe Reaction Mix, 1 μL of MPXV-B7 Forward Primer (10μM), 1μL of MPXV-B7 Reverse Primer (10μM), 0.5 μL of MPXV-B7 Taqman probe (10μM) with FAM as the reporter dye and BHQ1 as quencher, 2.5μL of Nuclease Free water and 5μL of DNA template. The PCR with recording of fluorescence intensity was performed according to the following protocol: 10 min at 94 °C and 40 cycles of 30s at 95 °C and 30s at 60°C.

For positive control, double stranded oligonucleotides containing target fragments of the B7 gene were synthesized (Macrogen, USA). The target fragment was subsequently cloned using TA cloning (PGMT Easy-Vector system, Promega, USA) to generate the plasmid pMPXV-B7. The plasmid pMPXV-B7 was then used as a positive control and to estimate copy number of MPXV DNA.

The copy number of MPXV DNA was estimated based on a standard curve generated by using pMPXV-B7 in a series of 10-fold serial dilutions. The dilution series ranged between 2 × 10^6^ copies /μL and 2 × 10^3^ copies/μL. The threshold of positive detection was set at <40 quantitative cycles (Cq). The limit of detection of the assay, determined based on Minimum Information for Publication of Quantitative Real-Time PCR Experiments (MIQE) guidelines (Huggett, 2020), was approximately 87 copies per reaction.

### Virus culture

Virus culture was performed for selected MPXV DNA positive samples. Briefly, 100 μL of the processed sample was diluted in 900 μL M199 media and passed through a 0.45 μM filter. Approximately 500 μL of the filtrate was inoculated on monolayer African Green Monkey kidney cells (Vero cells) grown in T-25 cell culture flasks. The inoculum was incubated with the cells for 1 hour at 37°C and 5% CO_2_ with gentle rocking for 15 secs every 15 mins before the addition of media. The flasks were then incubated and observed for cytopathic effect (CPE). Positive virus cultures were confirmed by qPCR. Samples were considered to be negative for virus culture after two subsequent passages with no CPE observed and with the cell culture extracts tested to be negative for MPXV DNA by qPCR.

### Result analysis

Graphpad Prism was used to carry out statistical analyses.

## Results

We sampled the hospital room environment occupied by a male patient admitted with skin lesions and an episode of fever. He had no other symptoms. MPXV DNA was detected from a nasopharyngeal swab and swab of peri-anal lesions on the day of admission (Day 5 of illness). Most of his skin lesions were located on the buttocks (n=23 lesions), followed by the back (n=15 lesions) and extremities (n=2-4 lesions), with no new lesions after day 8 of illness. The clinical course was uneventful, and the patient was discharged on day 23. We collected 179 environmental samples (air, n=56; surface, n=100; dust, n=16; water, n=7) on days 7, 8, 13, 15, and 21 of his illness. Details of the sampling schedule and the number of samples collected are provided in Supplementary Tables 1 and 2.

### Environmental contamination trend

Air contamination was highest during week 1 of illness (up to 11 copies/L air), with a gradual decline over the weeks to less than 1 copies/L of air by day 21 (Figure 1.A and 1.B). Similarly, surface contamination peaked on day 8 of illness (1.25 × 10^4^ copies/swab) and declined to the lowest contamination level by day 21 of illness (Figure 1.C). A similar trend was observed for vacuumed dust contamination, with the highest viral load observed on day 7 of illness (5.94 × 10^7^ virus copies/sample) and lowest on day 21 of illness (2.83 × 10^3^ to 2.29 × 10^5^ virus copies/sample) (Figure 1.D). Viable virus was isolated from four surface and dust samples.

**Figure 1.**
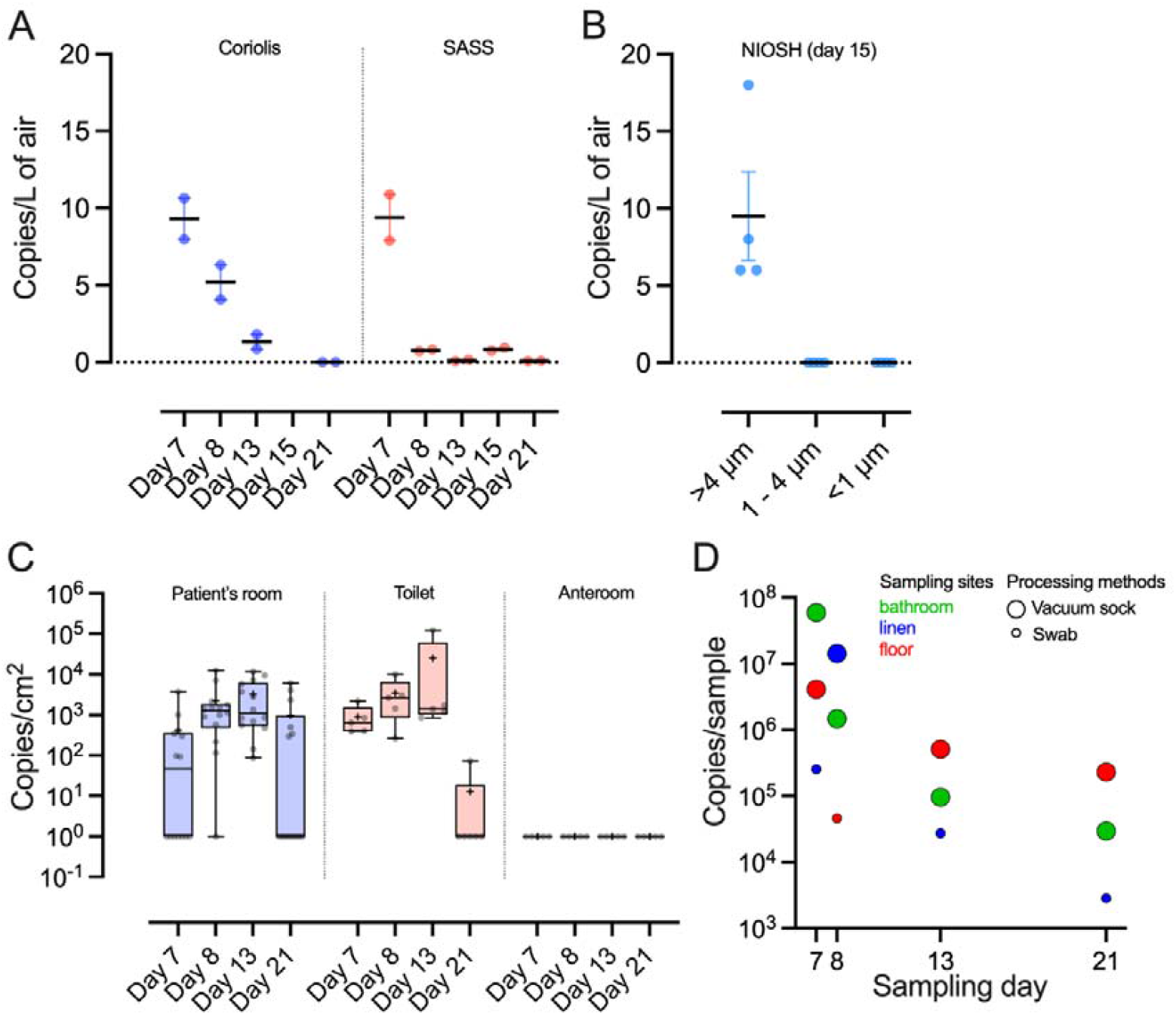
Environmental Contamination of Monkeypox Virus DNA. Virus DNA concentration (copies/L) detected in air using (A) Coriolis or SASS samplers across sampling days, and (B) NIOSH sampler on day 15 of disease. Line and error bars indicate mean and standard error of the mean (SEM). (C) Virus DNA concentration (copies/cm2) from swabs of environmental surfaces of patient room, toilet and anteroom across sampling days. Bar indicates median, + indicates mean. Box encompasses the interquartile range, and whiskers show the minimum to maximum values. (D) Virus DNA concentration (copies/sample) from dust samples across sampling dates.

### Air samples

The room was designed to be ventilated at 12 air changes per hour. MPXV DNA concentration in the air samples ranged from less than 1 to 11 copies per L of air sampled. Viral DNA was detected in all air samples from SASS samplers deployed at air flow rate of 300 L/min within 1 meter from the patient (Table 1). SASS samplers located at 2.5 meters from the patient detected MPXV DNA on days 13 and 15 but not on day 21. MPXV DNA concentration decreased from day 7 to 21. Similarly, Coriolis air samplers (air flow rate of 100 L/min) set within 1 meter of the patient detected MPXV DNA, decreasing concentration during the sampling period (days 7, 8, and 13). On day 15 of illness, all 4 NIOSH TE-BC251 air samplers deployed detected MPXV DNA in a particle-size fraction of > 4 μm, with 2-6 copies of MPXV DNA per L of air sampled. Using DustTrakTM DRX 8534 to monitor the particle sizes in the air on day 7 and 8, we observed no significant change (p> 0.05) in the concentration of particle matter of > 4 μm (size range at which we detected MPXV genetic material) before and after the air samplers were turn on (supplementary figure 2), but increased number of airborne particles during activities such as opening of doors (supplementary figure 3). No viable virus was isolated from any of the air samples.

**Table 1:** Monkeypox virus detection in the air of the room occupied by the infected patient

| Days of illness | Symptoms reported by patient | Patient's activities during sampling | Sampler type | Distance from patient's head (meter) | Aerosol particle size | Average Gene Copies/ PCR Reaction | Airborne Monkeypox virus concentration (copies/L of air) | Virus Culture |
| --- | --- | --- | --- | --- | --- | --- | --- | --- |
| Day 7 | Formation of new vesicles | In bed throughout sampling. Conversing over the phone | SASS<sup>R</sup> | 0.9 | NA | 6538 | 11 | - |
|  |  |  | SASS<sup>L</sup> | 0.8 | NA | 4743 | 8 | - |
|  |  |  | Coriolis<sup>R</sup> | 0.9 | NA | 639 | 11 | Negative |
|  |  |  | Coriolis<sup>L</sup> | 0.8 | NA | 478 | 8 | Negative |
| Day 8 | Formation of new vesicles | In bed throughout sampling. Conversing over the phone | SASS<sup>R</sup> | 0.9 | NA | 492 | 1 | - |
|  |  |  | SASS<sup>L</sup> | 0.8 | NA | 430 | 1 | - |
|  |  |  | Coriolis<sup>R</sup> | 0.9 | NA | 189 | 6 | - |
|  |  |  | Coriolis<sup>L</sup> | 0.8 | NA | 245 | 4 | Negative |
| Day 13 | No new vesicles. Some scabs seen | In bed throughout sampling. Conversing over the phone | SASS<sup>R</sup> | 0.9 | NA | 36 | <1 | - |
|  |  |  | SASS<sup>L</sup> | 0.8 | NA | 99 | <1 | - |
|  |  |  | SASS-F | 2.5 | NA | 56 | <1 | - |
|  |  |  | Coriolis<sup>R</sup> | 0.9 | NA | 54 | 2 | - |
|  |  |  | Coriolis<sup>L</sup> | 0.8 | NA | 46 | 1 | Negative |
|  |  |  | Coriolis-F | 2.5 | NA | ND | ND | - |
| Day 15 | No new vesicles. More scabs seen | - Around 2 hours were spent sitting on chair<br>- Around 1 hour was spent in bed<br>- Around 1 hour spent on exercise and walking around the room (Exercise on Yoga mat during first 30 mins and last 30 mins sampling) | SASS<sup>R</sup> | 0.9 | NA | 562 | 1 | - |
|  |  |  | SASS<sup>L</sup> | 0.8 | NA | 450 | 1 | - |
|  |  |  | SASS-F | 2.5 | NA | 344 | 1 | - |
|  |  |  | NIOSH<sup>R-1</sup> | 0.9 | > 4 µm | 88 | 6 | Negative |
|  |  |  |  |  | 1 -4 µm | ND | ND | - |
|  |  |  |  |  | < 1 µm | ND | ND | - |
|  |  |  | NIOSH<sup>R-2</sup> | 0.9 | > 4 µm | 31 | 2 | Negative |
|  |  |  |  |  | 1 -4 µm | ND | ND | - |
|  |  |  |  |  | < 1 µm | ND | ND | - |
|  |  |  | NIOSH<sup>L-1</sup> | 0.8 | > 4 µm | 40 | 3 | Negative |
|  |  |  |  |  | 1 -4 µm | ND | ND | - |
|  |  |  |  |  | < 1 µm | ND | ND | - |
|  |  |  | NIOSH<sup>L-2</sup> | 0.8 | > 4 µm | 29 | 2 | Negative |
|  |  |  |  |  | 1 -4 µm | ND | ND | - |
|  |  |  |  |  | < 1 µm | ND | ND | - |
| Day 21 |  |  | SASS<sup>R</sup> | 0.9 | NA | 44 | <1 | - |
|  |  |  | SASS<sup>L</sup> | 0.8 | NA | 53 | <1 | - |
|  |  |  | SASS-F | 2.5 | NA | ND | ND | - |
|  |  |  | Coriolis<sup>R</sup> | 0.9 | NA | ND | ND | - |
|  |  |  | Coriolis<sup>L</sup> | 0.8 | NA | ND | ND | - |

<sup>R</sup>Right side of the patient, <sup>L</sup>Left side of the patient, <sup>R-1</sup>First sampler on the right side, <sup>R-2</sup>Second sampler on the right side, <sup>L-1</sup>First sampler on the left side, <sup>L-2</sup>Second sampler on the left side
Dashes ‘-’, not done; NA, not applicable; ND, not detected; All controls were negative.

#### Surface samples

Surface sampling was conducted before the daily cleaning and disinfection of the room. Out of the 19 samples collected from the room and toilet each day, most were positive MPXV DNA, with the number of positive samples increasing from 12 on day 7 to 18 on day 8 and day 13, before subsiding to 7 on day 21 (Table 2). The highest viral load (virus gene copies/cm^2^) was detected on the patient bathroom’s internal sink surface, patient’s room floor and call bell. All samples from the anteroom and clean corridor were negative for MPXV DNA. Viable virus was isolated using VERO cells from the toilet seat (day 7 of illness) and the chair in the room (day 8 of illness).

**Table 2:**
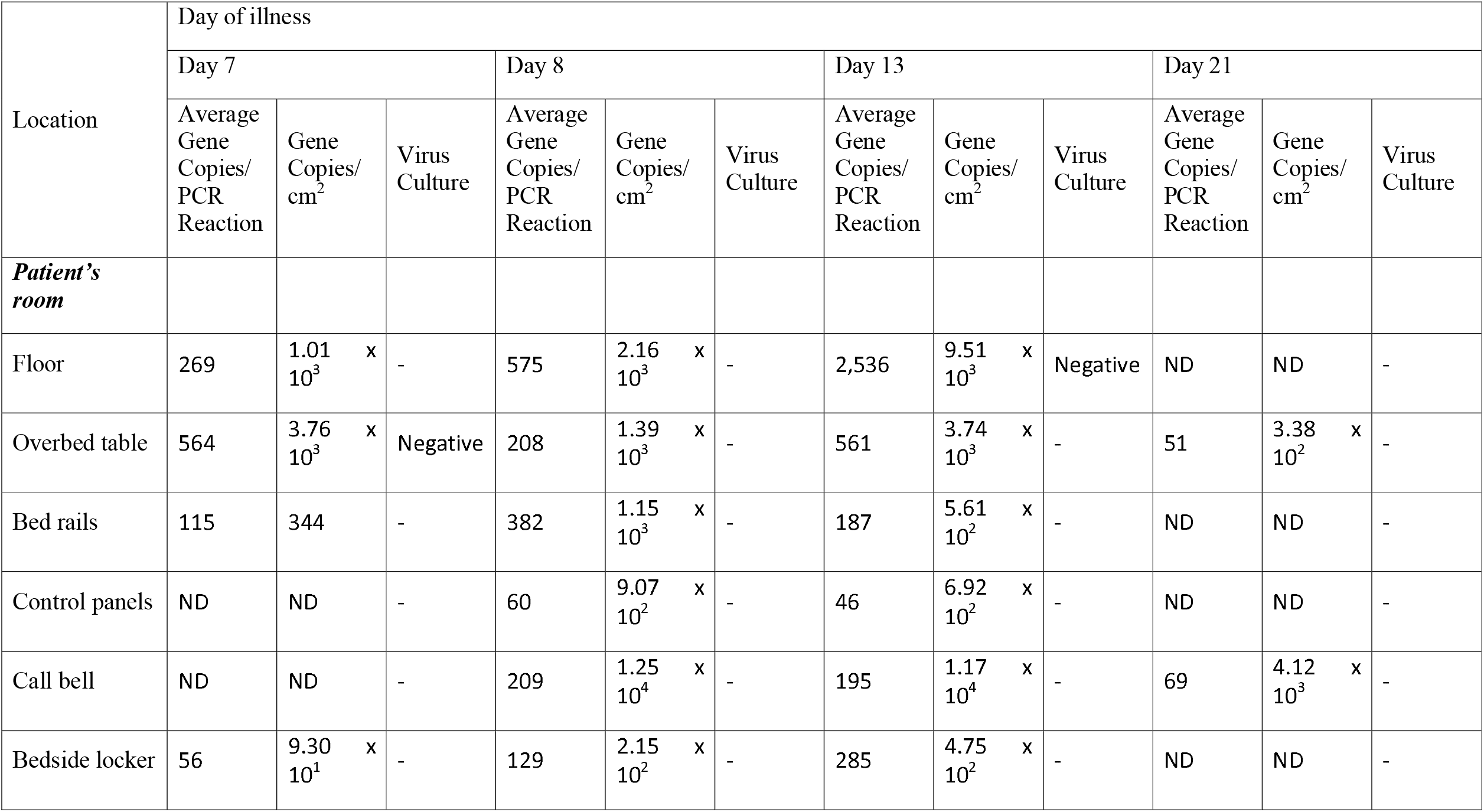
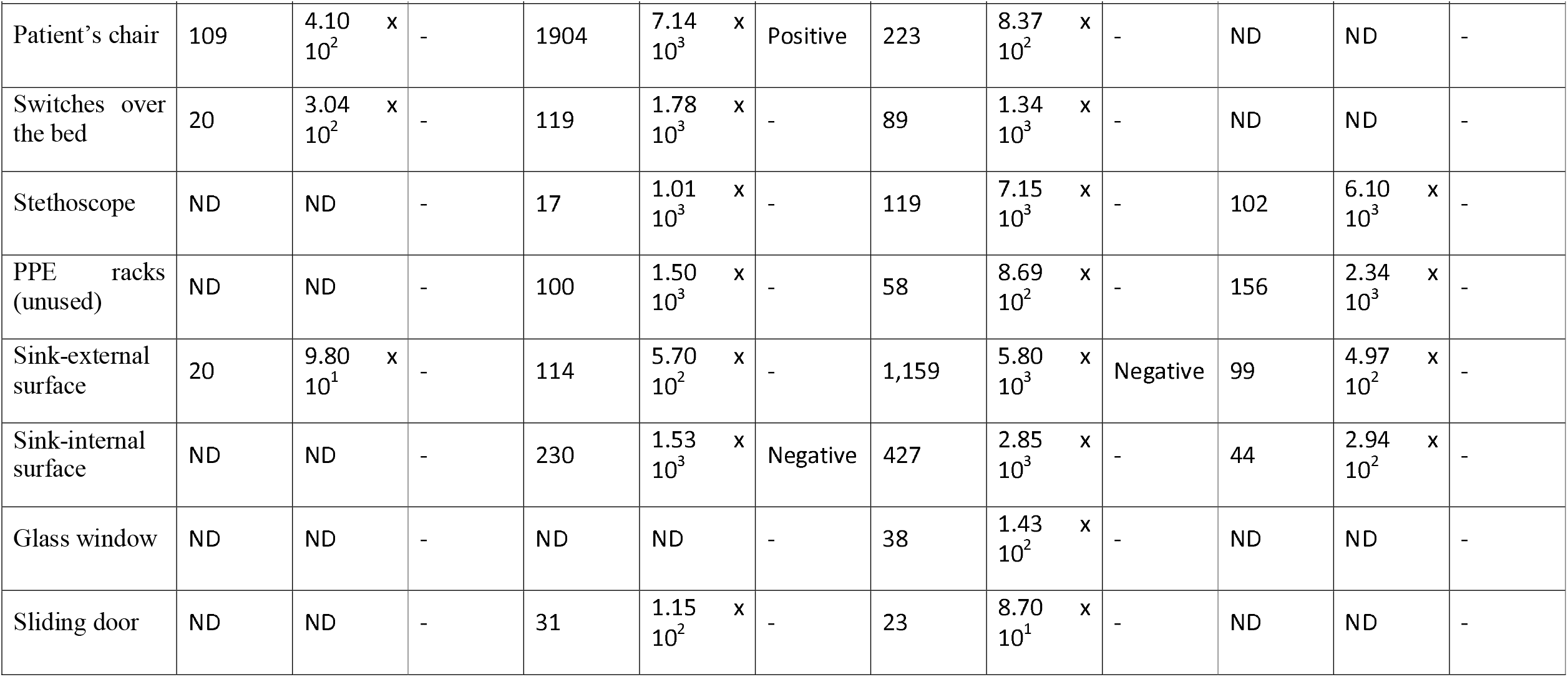
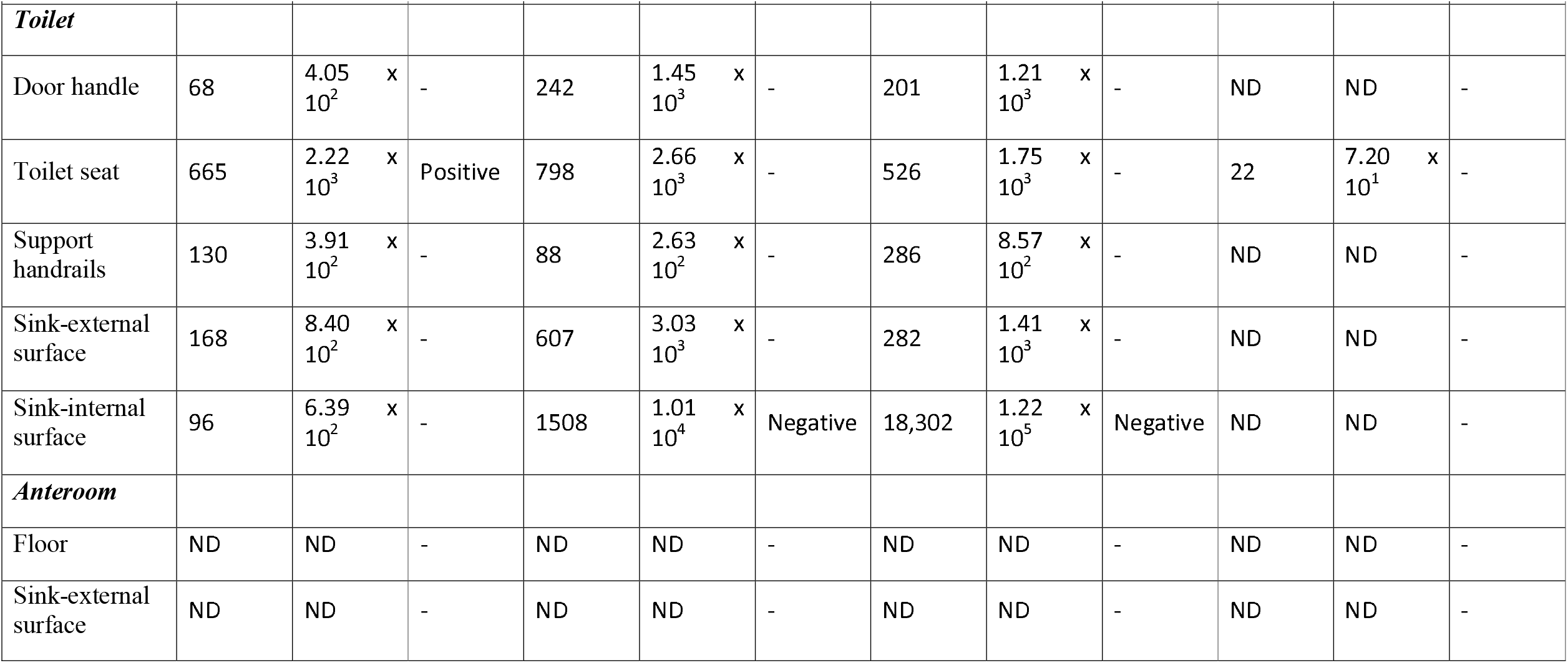
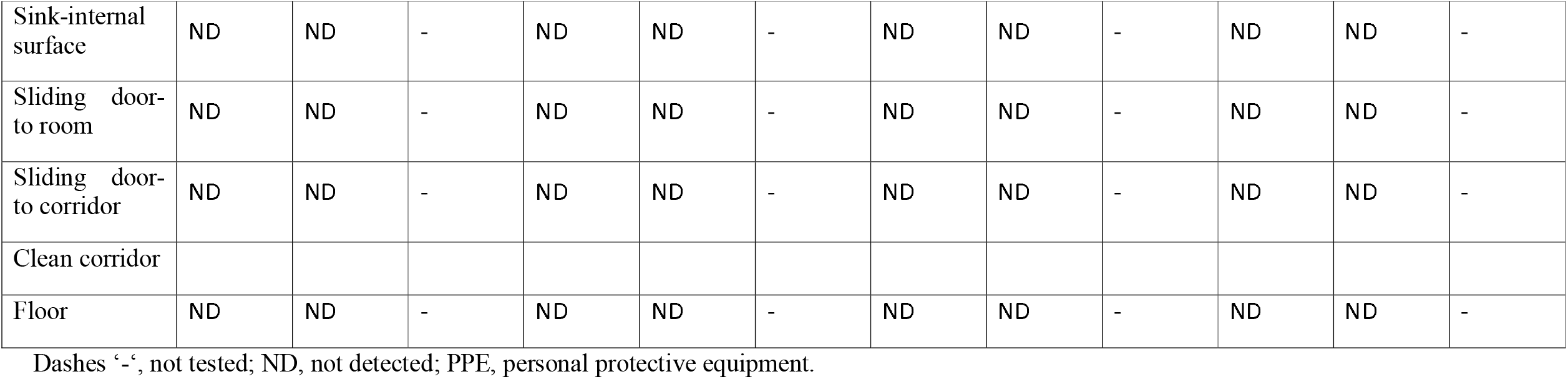
Monkeypox virus detection on surfaces of the room occupied by an infected patient

#### Dust and water samples

Dust samples from linen and floors of room and toilet were collected using vacuum socks (X-Cell-200, Midwest Filtration) fitted on a vacuum pump (Omega Plus HEPA Abatement Vacuum, Atrix international), and MPXV DNA was detected in these samples until day 21. The highest viral load of all environmental samples was observed in toilet floor dust samples (5.94 × 10^7^ virus copies/sample) (Table 3). Viable virus was isolated from linen dust samples on days 7 and 8 of illness. Water from the Sink P-traps was positive for MPXV DNA until day 13 but became negative afterwards.

**Table 3:**
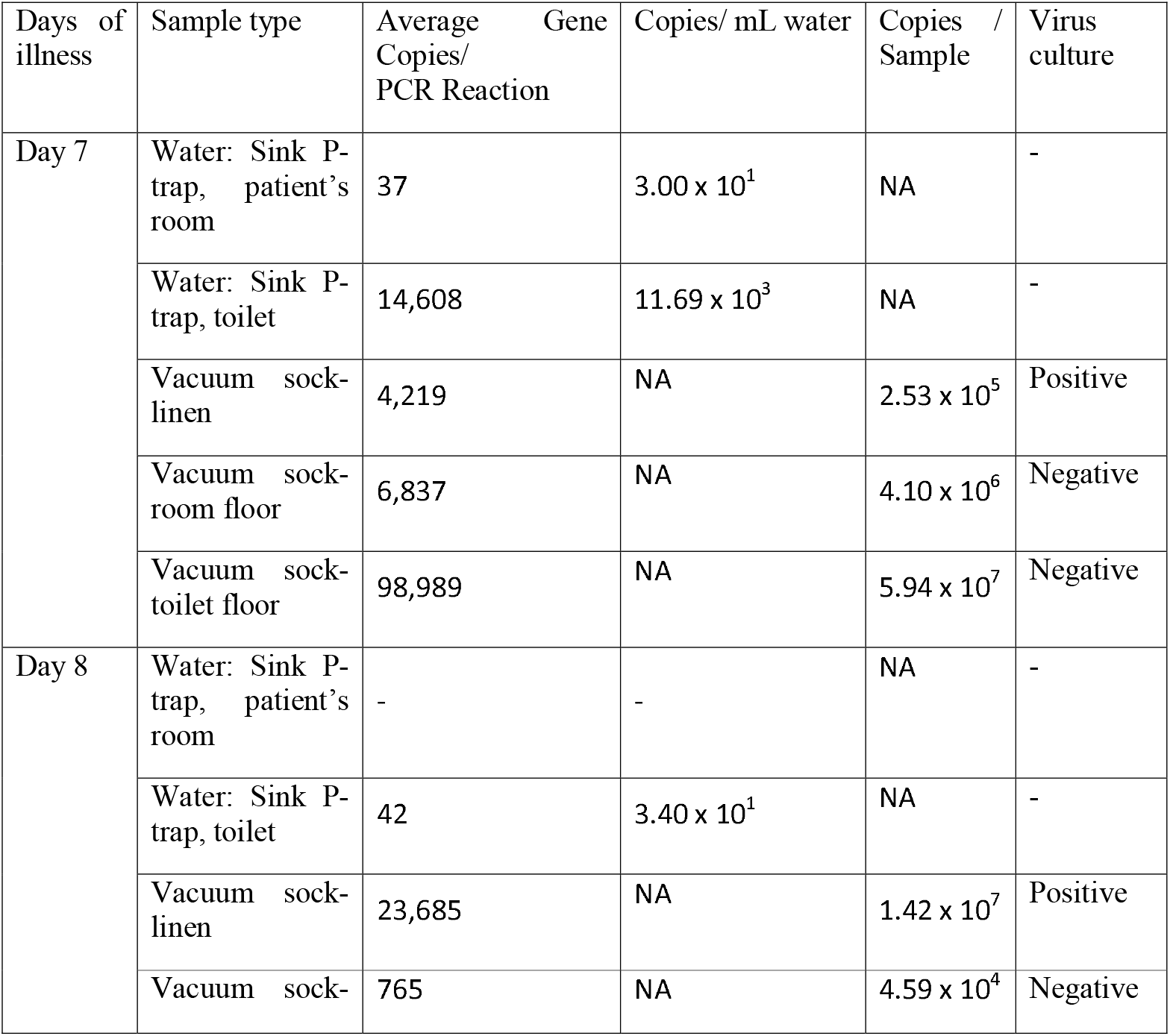
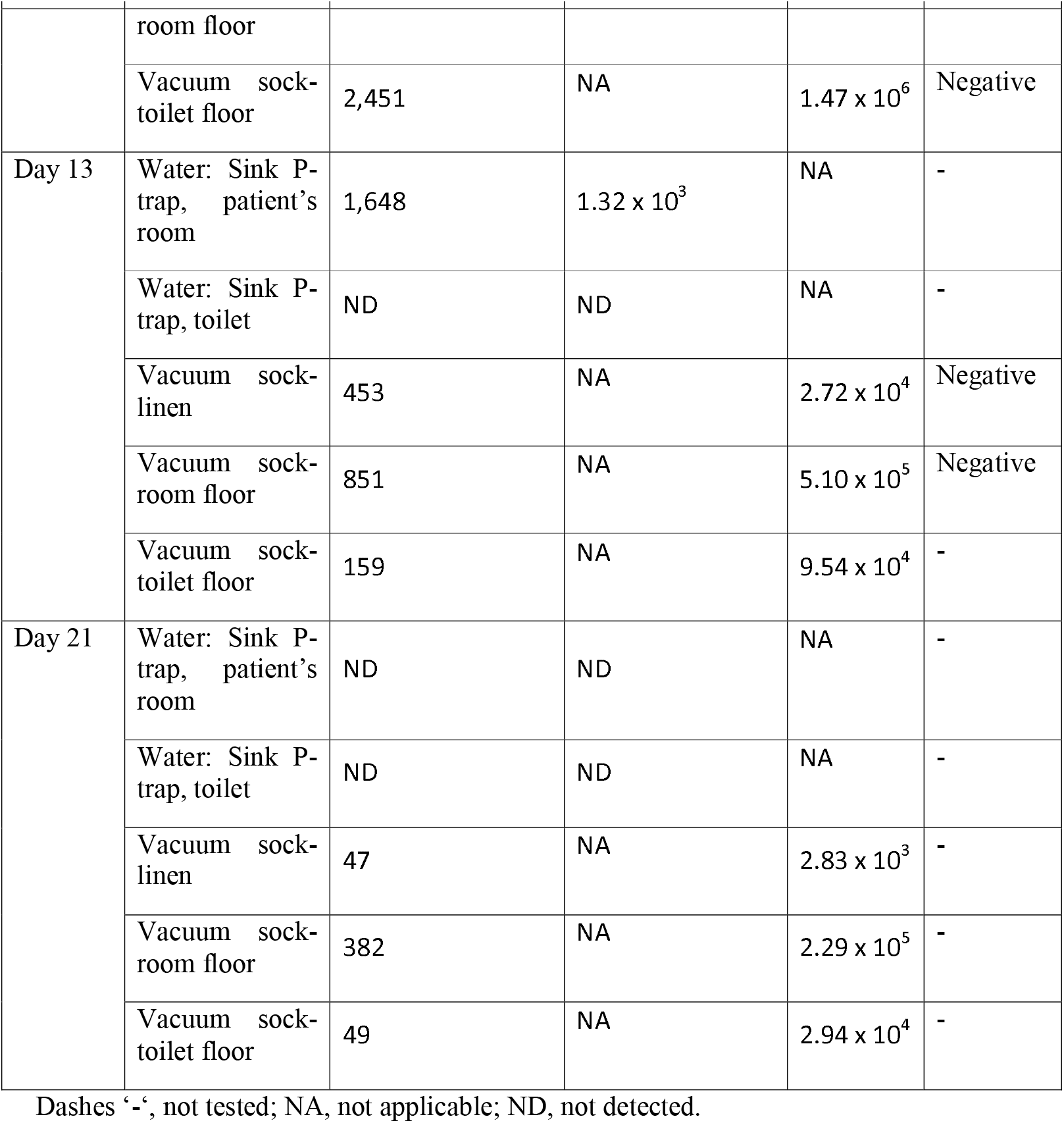
Monkeypox virus detection in water and vacuumed dust sample

## Discussion

In this study, swab and dust sampling showed extensive surface contamination of the hospital room occupied by a MPXV patient with the recovery of viable MPXV virus from the chair, toilet seat and dust from bed linen. MPXV DNA was consistently detected in almost all air samplers, though they were not culturable. Surface, dust, and air contamination gradually declined after the first week of illness.

The recovery of viable virus from the chair and toilet seat correlates with the location of skin lesions of the study patient. As linens were changed daily and surfaces and floors of the room and toilet were cleaned daily, the detection of MPXV DNA across sampling days showed continued viral shedding over the course of the illness. While environmental contamination has been reported in other recently published reports^7-9^, our study tracked virus shedding of the patient over time, with reduced levels of environmental contamination from the second week of illness onwards, coinciding with the period when the patient stops developing new lesions. Findings of viable viruses from surface swabs and dust samples support the possibility of fomite-based transmission, especially in the nosocomial^4^ and home settings. It highlights the importance of surface disinfection, especially of chairs, toilets and floors, and the need for precautions when handling linens.

The daily detection of viral particles by all air samplers in an environment with 12 HEPA-filtered unidirectional air changes per hour, together with previous findings of viable airborne MPXV virus in the United Kingdom^9^, underscores the possibility of aerosol-based transmission of MPXV. Our finding of viral material only in particles of > 4 μm sizes suggests that the possibility of breathing and/or talking being the source of the virus is low, as these activities were previously found to emit predominantly virus particles of <5 um sizes^10, 11^. However, studies in a more typical environment without such high ventilation rates or more direct sampling of the breath is needed to verify and provide a better understanding of respiratory, talking and/or coughing source of the viral particles. On the other hand, the presence of viruses, including live virus, in dust samples suggests lesion shedding as the potential source of contaminated particles in the air. As demonstrated by the temporal number of particles of various sizes, the number of contaminated airborne particles could be influenced by activities that impact flow current in the space, such as opening doors and presumably changing linens.

Notably, the detection of MPXV DNA from wastewater in the sink traps provides direct evidence for the possible utility of wastewater-based surveillance^1^ of MPXV. One limitation of this study is that we analyzed only one patient, and the level of environmental contamination may differ for patients who may not have as extensive skin lesions (e.g. proctitis only) or who are asymptomatic. Further data is needed for these scenarios. The finding of extensive environmental contamination and the presence of detectable MPXV DNA in the air must be considered in the larger context of other factors in assessing the transmission dynamics of MPXV, including the infectious dose required for manifestation of the disease. The potential for onward transmission is affected at least by the inoculum dose and host susceptibility for any particular transmission mode. Some have suggested that pathogens deemed capable of aerosol transmission should be associated with high R0. However, current practice and the evidence do not support this in all instances. For example, pertussis, a pathogen traditionally considered a droplet transmitted pathogen, has a much higher R0 than tuberculosis, a pathogen transmitted via aerosols^12^. As such, in the background of a steadily increasing global outbreak and the natural ability of MPXV to mutate, all possible modes of transmission should be addressed when considering transmission prevention, especially in the healthcare setting.

## Data Availability

All data produced in the present work are contained in the manuscript

## Associated content

### Supporting Information

Supplementary figures and tables are found in the Supplementary material file.

## Acknowledgements

We would like to thank the Ministry of Health, Singapore for their review of the manuscript. We also appreciate the nursing team at the National Centre of Infectious Diseases, and the Facilities, Safety and Quality Department at the Environmental Health Institute, for their support in sample collection and coordination. We would like to thank Dr Sean Ong for the graphics and Mr Pek Han Bin for his support in sample transfer.

## Author Contributions

The manuscript was written through contributions of all authors. All authors have given approval to the final version of the manuscript. ^#^These authors contributed equally.

## Ethics statement

Informed consent was obtained from the patient as part of the PROTECT study protocol (DSRB number: 2012/00917).

This study was approved by the Environmental Health Institute’s Management Committee (Project TS325), National Environment Agency.

