## Supplementary Materials for "Viable Monkeypox virus in the environment of a patient room"

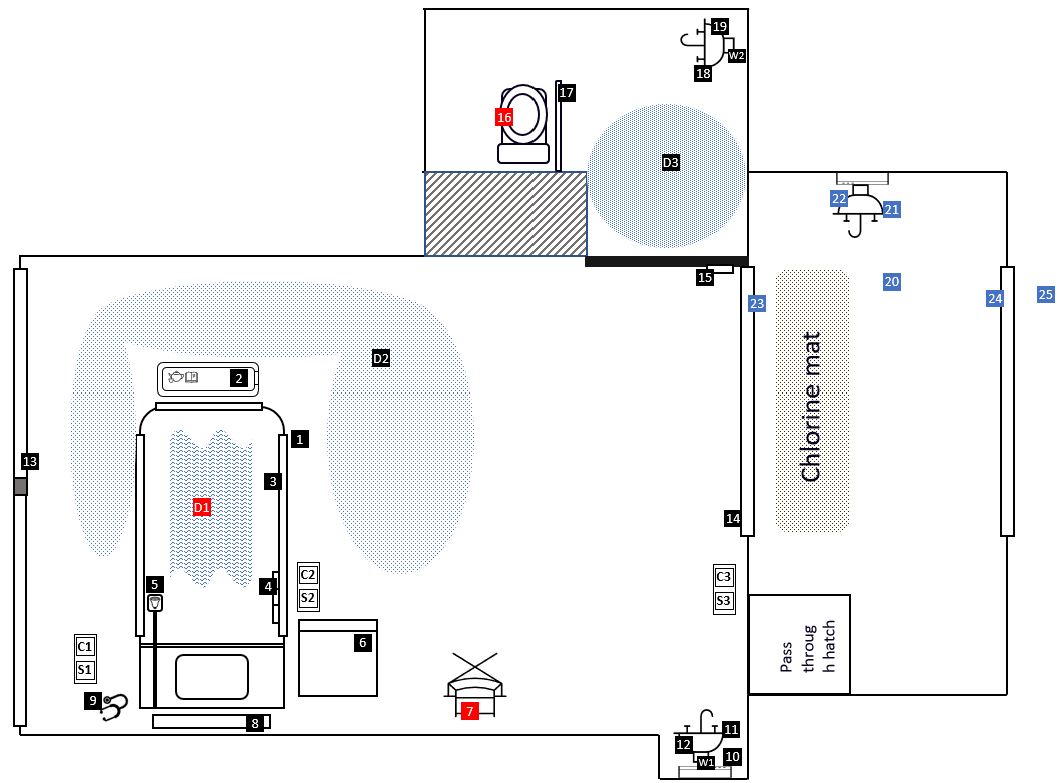

**Supplementary Figure 1. Room Layout and sampling locations**

*Patient room:* (1) Patient room floor, (2) Overbed tables, (3) Bed rails, (4) Bed control panels, (5) Call bell, (6) Bedside locker, (7) Patient’s chair, (8) Switches over the bed, (9) Stethoscope, (10) Personal protective equipment rack, (11) Sink-external surface, (12) Sink-internal surface, (13) Glass window, (14) Sliding door, *Toilet:* (15) Toilet door handle, (16) Toilet seat, (17) Support handrails, (18) Toilet sink-external surface, (19) Toilet sink-internal surface, *Anteroom:* (20) Floor, (21) Sink-external surface, (22) Sink-internal surface, (23) Sliding door to patient room, (24) Sliding door to clean corridor, *Clean corridor:* (25) Floor. W1, water from room sink P-trap; W2, water from toilet sink P-trap; D1, vacuumed dust sample from linen; D2, vacuumed dust sample from patient room floor; D3, vacuumed dust sample from toilet floor. For D1, D2, and D3, the corresponding shaded area represents the vacuumed area. C1, Left Coriolis air sampler (0.8 meters from patient); S1, Left SASS air sampler (0.8 meters from patient); C2, right Coriolis air sampler (0.9 meters from patient); S2, right SASS air sampler (0.9 meters from patient); C3, Coriolis air sampler at 2.5 meters from patient; S3, SASS air sampler at 2.5 meters from the patient.

Red color marks surfaces with viable Monkeypox virus; Blue color mark samples that were never positive for Monkeypox virus DNA.

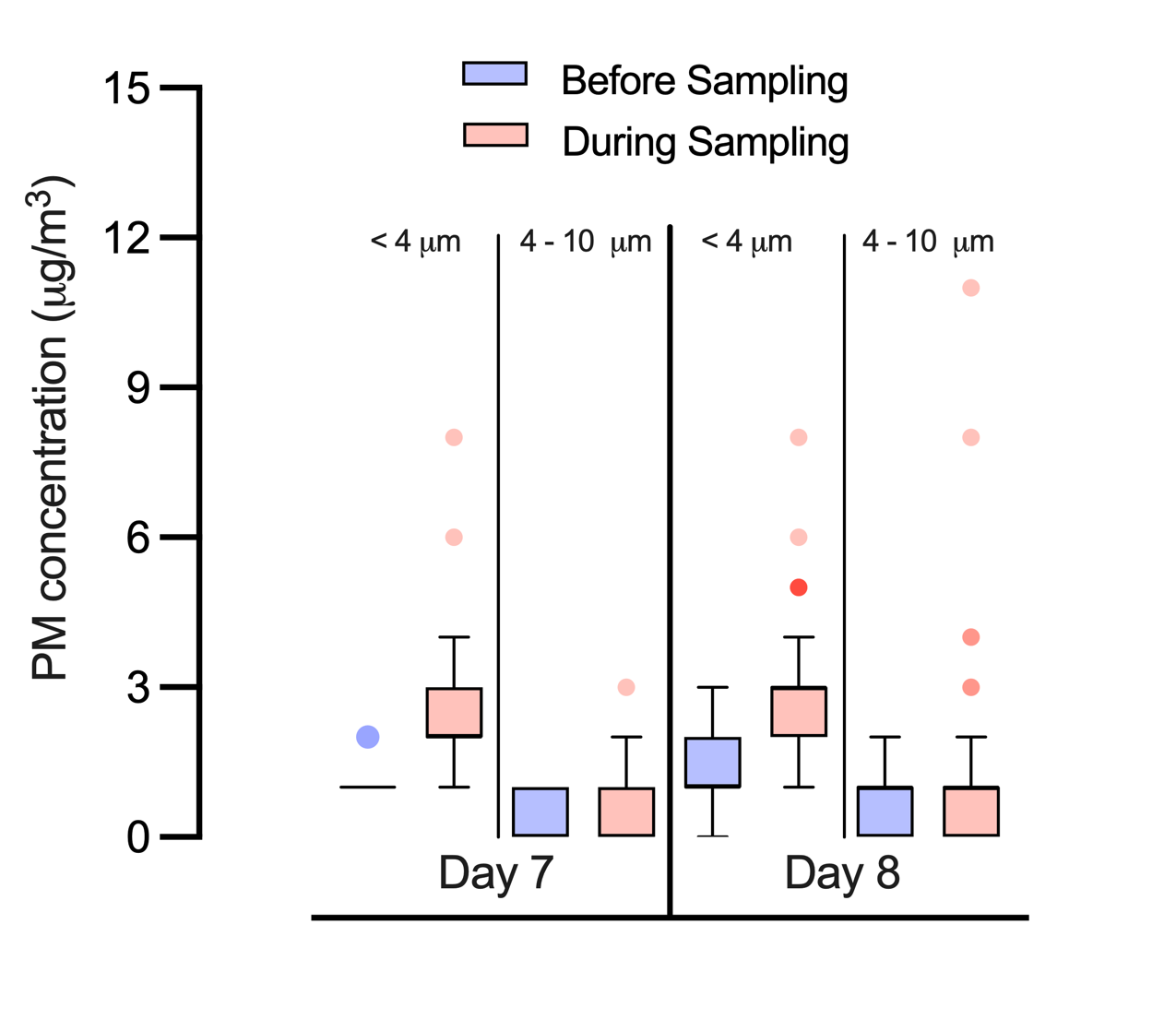

**Supplementary Figure 2. Mass concentration of particulate matter (PM) <4µm and 4-10µm before and during sampling on day 7 and 8 of illness.**

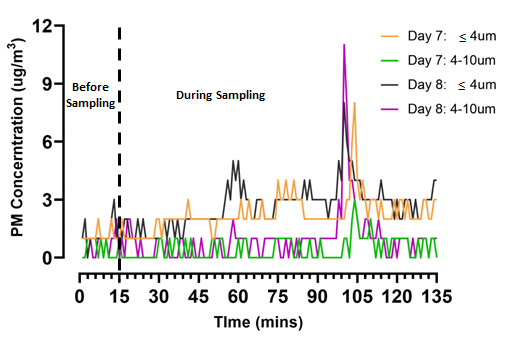

**Supplementary Figure 3. Time-of particulate matter (PM) <4µm and 4-10µm concentration before and during air sampling on day 7 and 8 of illness.** Sampling personnel exiting the room at around the 96 minute mark may have led to an spike in particles concentration.

**Supplementary Table 1. Environmental sampling schedule**

| **Sample types** | **Day 6** | **Day 8** | **Day 13** | **Day 15** | **Day 21** |
| --- | --- | --- | --- | --- | --- |
| Air samples* |  |  |  |  |  |
| SASS3100 | 2 units | 2 units | 3 units | 3 units | 3 units |
| Coriolis µ | 2 units | 2 units | 3 units | No | 3 units |
| DustTrak^TM^ DRX 8534 | 1 unit | 1 unit | 1 unit | No | 1 unit |
| NIOSH BC 251 | No | No | No | 4 units | No |
| Surface samples | Yes | Yes | Yes | No | Yes |
| Dust (Vacuumed samples) |  |  |  |  |  |
| Linen | Yes | Yes | Yes | No | Yes |
| Room floor | Yes | Yes | Yes | No | Yes |
| Toilet floor | Yes | Yes | Yes | No | Yes |
| Water samples |  |  |  |  |  |
| P-trap (room) | Yes | Yes | Yes | No | Yes |
| P-trap (toilet) | Yes | Yes | Yes | No | Yes |

*Please refer to Figure 1 for the location of air samplers in the room

**Supplementary Table 2. Total number of samples collected**

| **Sample types** | **Number of samples by day of illness** | | | | | **Total** |
| --- | --- | --- | --- | --- | --- | --- |
|  | **Day 7** | **Day 8** | **Day 13** | **Day 15** | **Day 21** |  |
| **Air** |  |  |  |  |  |  |
| SASS3100 | 4 | 4 | 6 | 6 | 6 | 26 |
| Coriolis µ | 4 | 4 | 6 |  | 4 | 18 |
| NIOSH BC 251 |  |  |  | 12^1^ |  | 12^1^ |
| *Subtotal* | 8 | 8 | 12 | 18 | 10 | 56 |
| **Surface** |  |  |  |  |  |  |
| Room | 14 | 14 | 14 |  | 14 | 56 |
| Toilet | 5 | 5 | 5 |  | 5 | 20 |
| Anteroom | 5 | 5 | 5 |  | 5 | 20 |
| Clean corridor | 1 | 1 | 1 |  | 1 | 4 |
| *Subtotal* | 25 | 25 | 25 | 0 | 25 | 100 |
| **Dust** |  |  |  |  |  |  |
| Linen | 1 | 1 | 1 |  | 1 | 4 |
| Room floor | 1 | 1 | 1 |  | 1 | 4 |
| Toilet floor | 1 | 1 | 1 |  | 1 | 4 |
| Control | 1 | 1 | 1 |  | 1 | 4 |
| *Subtotal* | 4 | 4 | 4 | 0 | 4 | 16 |
| **Water** |  |  |  |  |  |  |
| Toilet sink P-trap | 1 | 1 | 1 |  | 1 | 4 |
| Room sink P-trap | 1 |  | 1 |  | 1 | 3 |
| *Subtotal* | 2 | 1 | 2 | 0 | 2 | 7 |
| **Total number of samples** | **39** | **38** | **43** | **18** | **41** | **179** |
